# In Utero Infection with Coxsackievirus-B is Associated with Congenital Pulmonary Atresia

**DOI:** 10.1101/2020.04.18.20070813

**Authors:** Vipul Sharma, Lisa S. Goessling, Anoop K. Brar, Pirooz Eghtesady

## Abstract

The research for this study was undertaken to examine association between perinatal infection with Coxsackievirus B (CVB) and congenital heart defects (CHD). Suggestive, although inconclusive, data exists of an association between CVB infection during pregnancy and CHD. We present data from a clinical study showing that CVB infection in early pregnancy induces heart defects. In a prospective study of 123 pregnant women with pregnancies affected with CHD we found circulating CVB IgG and IgM in the serum. A significant number of the women were positive for CVB antibodies; 30.9% for IgG and 6.5% for IgM. Our data demonstrates not only an association of CVB with CHD in pregnant women but this is the first report showing significant association between in-utero CVB infection and Pulmonary Atresia (PA) and HLHS (AA/MS variant). Elevated levels of IgG were seen in 55.5% of women carrying babies with PA and 60% of women carrying babies with HLHS (AA/MS), with p-values of 0.01 and 0.03 respectively. To determine specific CVB serotypes involved, neutralizing antibody titers to CVB1, CVB3, and CVB4 were analyzed on samples with positive IgG/IgM levels, and showed a possible association between CVB4 and CHD in the cohort. Our study has broad implications for the future understanding and potential management of CHD as well as introduces a new avenue of research for cardiac defects.

## Introduction

Large epidemiologic studies done in the early 1960s suggested an association between maternal Coxsackievirus-B (CVB) infection and congenital heart defects (CHD) (1), however, no association with specific defects was identified at the time. Using a murine model, our lab has shown that maternal CVB infection during gestation can induce fetal CHD, mostly Ventricular Septal Defect and Non-Compaction of ventricular myocardium (2). To investigate the potential clinical relevance of these findings, we conducted a prospective study of pregnant women carrying babies affected by CHD recruited over a 5-year period. Maternal Serum Coxsackievirus IgG and IgM levels were measured to determine if there is an association between maternal CVB exposure and development of CHD. To determine specific CVB serotypes involved, neutralizing antibody titers to CVB1, CVB3, and CVB4 were analyzed on samples with positive IgG/IgM levels.

## Methods

A total of 123 pregnant women referred for fetal echocardiography, due to concern for possible CHD, were enrolled and followed to term to confirm the presence and type of CHD in the baby. The study was approved by Washington University School of Medicine Institutional Review Board under protocols 201102410 and 201602122. Detection of CVB IgM and IgG antibodies was performed using Serion ELISA classic kits (ESR134M and ESR134G, QED Biosciences) as per manufacturer’s instructions. Titers of 15U/ml and above were considered positive. Neutralizing antibodies titers were measured against CVB1-Chi07 (provided by Dr. Xiaotian Zheng with Ann & Robert H. Lurie Children’s Hospital of Chicago), CVB3-Nancy, and CVB4-J.V.B. (ATCC). An antibody titer of 80 and above was considered elevated.

## Results

### Evidence of CVB exposure/infection in pregnant women

A positive IgG or IgM antibody level was considered an indicator of possible in-utero CVB exposure/infection. A significant number of the women in our study were positive for CVB antibodies; 30.9% for IgG and 6.5% for IgM.

### In-utero CVB exposure/infection is significantly associated with Pulmonary Atresia and HLHS (AA/MS)

The types of CHD present in pregnancies showing positive levels of CVB IgG and IgM were examined. Among the numerous types of CHD seen in our cohort, Pulmonary Atresia (PA) and Hypoplastic Left Heart Syndrome with Aortic Atresia/Mitral Stenosis variant (HLHS AA/MS) were found to be significantly associated with elevated CVB IgG. Elevated levels of IgG were seen in 55.5% (10/18) of women carrying babies with PA and 60% (6/10) of women carrying babies with HLHS (AA/MS), with p-values of 0.01 and 0.03 respectively. Of the 8 women with positive IgM levels, 25% were carrying babies with PA (p-value 0.4).

### Possible association between CVB4 and CHD in the cohort

Among the IgG positive samples, the percentage of elevated titers were 28.9% (11/38) to CVB1, 52.6% to CVB3 (20/38), and 68.4% to CVB4 (26/38), with 39.5% (15/38) samples having elevated titers to more than one serotype. Of the 10 PA affected, CVB IgG positive pregnancies, 40% had anti-CVB1 titers, 70% had anti-CVB3 titers, and 50% had anti-CVB4 titers, with 50% having elevated titers to more than one serotype. Of the 6 HLHS (AA/MS), IgG positive cases, none had elevated anti-CVB1 titers, 50% had anti-CVB3 titers, and 83.3% had anti-CVB4 titers, with 20% having elevated titers to both CVB3 and CVB4. In contrast, all of the IgM positive samples had elevated titers to only CVB4, however no association with a specific CHD was seen.

## Discussion

This study shows a significant association between CVB in-utero infection/exposure and PA and HLHS AA/MS variant. Acquired PA has been associated with twin-to-twin transfusion syndrome, mechanism for which remains unknown (3). It is of interest that in both of these CHD, coronary sinusoids are a typical feature. Another interesting observation is one of the subjects with high CVB IgG had a fetus affected by isolated RV associated coronary fistula. Together, these findings suggest fetal exposure to CVB could perhaps lead to formation of abnormal myocardial architecture and associated sinusoids.

While interesting, our study has limitations. The sample group was biased by those specifically referred for fetal echocardiography and hence likely the more severe end of the CHD spectrum. Also, since study design only allowed enrollment later in pregnancy, only inferences can be made regarding earlier exposure based on antibody titers. A large prospective cohort study would be the only means to further assess the validity of our findings. Lastly, increased exposure risk to CVB (or susceptibility) maybe a surrogate for another factor such as diabetes that is actually causative for our observations. We welcome suggestions and inputs from other investigators to confirm these observations.

## Data Availability

The authors confirm that the data supporting the findings of this study are available within the article.

## References

1. Brown GC, Evans TN. Serologic evidence of Coxsackievirus etiology of congenital heart disease. Journal of the American Medical Association. 1967;199(3):183–7.

2. Sharma V, Goessling L, Brar A, Eghtesady P. Coxsackievirus infection during early pregnancy leads to congenital heart defects. Circulation. 2018;138:A16756.

3. Mieghem TV, Lewi L, Gucciardo L, et al. The fetal heart in twin-to-twin transfusion syndrome. International Journal of Pediatrics. 2010;2010:379792.

